# Predictors of HIV Testing among Youth 15-24 Years in urban Ethiopia, 2017-2018 Ethiopia Population-based HIV Impact Assessment

**DOI:** 10.1101/2022.03.17.22272053

**Authors:** Aderonke S. Ajiboye, Frehywot Eshetu, Sileshi Lulseged, Yimam Getaneh, Nadew Tademe, Tsigereda Kifle, Rachel Bray, Hailegnaw Eshete, Yohannes Demissie, Clare A. Dykewicz, David Hoos, EPHIA Study Group

## Abstract

**Introduction:** Youth (adolescents and young adults) aged 15-24 years comprise approximately 22% of Ethiopia’s total population and make up 0.73% of HIV cases in urban Ethiopia. However, only 63% of HIV-positive youth are aware of their HIV status. We described the HIV testing behaviors of youth 15-24 years and determined the characteristics of those who were most likely to be tested for HIV within the past year.

**Methods:** Using data from the 2017-2018 Ethiopia Population-based HIV Impact Assessment, we provide survey-weighted estimates and prevalence risk ratios for engagement in HIV testing in the 12 months preceding the survey. We model the likelihood of HIV testing one year or more before the survey compared to never testing, using a multinomial logistic regression model.

**Results:** Among HIV-negative and unaware HIV-positive youth 15-24 years old (N=7,508), 21.8% [95% Confidence Interval (CI): 20.4-23.3%] reported testing for HIV in the last 12 months. Female youth [Prevalence Ratio (PR)=1.6, 95% CI: 1.4-1.8], those aged 20-24 years (PR=2.6, 95% CI:2.3-2.9), and those ever married (PR=2.8, 95% CI: 2.5-3.1) were more likely to have tested for HIV within the last year. Adjusting for select demographic characteristics, sex with a non-spousal or non-live-in partner [Relative Risk (RR)=0.3, 95% CI:0.1-0.8] among males did not increase their likelihood to test for HIV in the prior 12 months. Female youth engaged in antenatal care (RR=3.0, 95% CI: 1.7-5.3) were more likely to test for HIV in the past year.

**Conclusion:** The Ethiopian HIV case finding strategy may consider approaches for reaching untested youth, with a specific focus on adolescent males. This is critical towards achieving the UNAIDS HIV testing goal of 95% of all individuals living with HIV aware of their status by 2030.

## INTRODUCTION

Since 1990, under-five child mortality in Eastern and Southern Africa has steadily decreased by 3.7% each year [1]. The reduction in child mortality over the last 30 years in sub-Saharan Africa has contributed to what is known today as the African ‘youth bulge’, with 60% of the total sub-Saharan African population under the age of 25 years [2]. The rapidly increasing young adult population across sub-Saharan Africa has introduced new challenges in the fight to end HIV/AIDS. Historically, perinatally infected children were assumed to not have survived past childhood. However, recent studies in Zimbabwe and South Africa have shown that an increasing number of undiagnosed adolescents are now presenting at primary care facilities with symptoms of long-term HIV infection [3] [4].

Many new cases of HIV infection are also occurring within the 15–24-year-old age group. As of 2019, 15–24-year-old sub-Saharan African youths make up 36% of all new HIV infections among adults 15 years and older [5]. In 2017 alone, an estimated 290,000 new HIV infections occurred among those 15-24 years in Eastern and Southern Africa with two-thirds of these infections among young women [2]. If not for the youth bulge, it is estimated that between 2010-2017, there would have been 340,000 fewer new cases of HIV among sub-Saharan African youth, aged 15-24 years [2]. As the youth population continues to grow across sub-Saharan Africa, the greatest challenge to reaching HIV epidemic control will be the identification of new and untreated cases of HIV.

In Ethiopia, Africa’s second most populous country, youths aged 15-24 years make up 22% of the nation’s total population [6]. In the urban and densely populated parts of the country, the prevalence of HIV is 4.2%, which is almost triple the national prevalence of 1.5% [6]. Little is known about how youth contribute to the national urban HIV epidemic in Ethiopia. The 2016 Ethiopia Demographic Health Survey assessed for knowledge of HIV transmission and prevention methods among youth. Survey results found that only 24% of women and 39% of men 15-24 years of age had a comprehensive knowledge of HIV prevention methods [7]. Prior to recent population-based estimates [8], little has been known of the actual burden of HIV in the younger age groups, specifically regarding overall HIV prevalence and the percentage of undiagnosed and untreated HIV infections.

The Ethiopia Population-based HIV Impact Assessment (EPHIA), conducted from 2017-2018, found that only 63% of HIV-positive 15–24-year-olds were aware of their HIV status. Among those who were aware, 100% were on antiretroviral HIV treatment, and 78.6% of those on treatment had achieved HIV viral load suppression [8]. Ethiopian HIV-positive youth are well below the first Joint United Nations Programme on HIV/AIDS (UNAIDS) recommended target of 90% awareness of HIV positive status [9]. The Ethiopia national HIV program has implemented several strategies to increase efforts to identify untreated cases of HIV. These strategies mainly employ provider-initiated case finding and social networking testing [10]. The services offered within these strategies, index case testing and partner notification, have historically targeted the married partners and biological children of HIV-positive individuals [10]. The current national HIV testing strategies may be missing many undiagnosed HIV-positive youths 15-24-years old in urban Ethiopia. This paper seeks to describe the HIV testing behaviors of Ethiopian youth 15-24 years, and to identify the demographic and behavioral characteristics of this age group, as well as the aspects of the healthcare system that facilitate the engagement of youth in HIV testing in urban Ethiopia.

## METHODS

The Ethiopia Population-based HIV Impact Assessment (EPHIA) was a national, household-based survey conducted in urban Ethiopia from October 2017 to March 2018. As a cross-sectional survey, the primary objective was to estimate the national and sub-national proportion of HIV viral load suppression (<1000 copies/mL) among HIV-positive adults 15-49 years of age. Secondly, the survey aimed to estimate the prevalence of HIV and the coverage of HIV prevention and treatment services among adolescents and adults aged 15-64 years. We assessed HIV testing behaviors of persons aged 15-24 years and focused on those who were HIV-negative or unaware of their HIV-positive status. We aimed to identify factors that might inhibit youth’s access to HIV testing, particularly among those at risk of acquiring HIV and those at risk of unknowingly transmitting the virus.

### Sampling design

The EPHIA used a two-stage cluster sampling design [11] to select a representative sample of enumeration areas (EAs) and households across urban areas of Ethiopia. Urban was defined as any region, district, or capital where residents do not engage in agricultural practices as the primary source of income. The 2007 National Ethiopian Census served as the sampling frame for the selection of enumeration areas. The first stage of sampling included 393 EAs selected randomly from all eleven regions of the country based on a probability proportional to regional population size. The second stage of sampling involved an average number of 30 households randomly selected per cluster, using an equal probability method, for a total of 11,810 households.

### Participant Eligibility and Data collection

A household questionnaire was administered to selected households. Eligible household members were those between the ages of 0-64 years old, those who slept in the household the night before the survey, spoke one of six survey languages and were willing and able to provide written informed consent or assent. Non-de facto household members, those who did not sleep in the household the night before the survey, were not eligible for participation. The questionnaire assessed for the biobehavioral factors related to HIV infection, including sociodemographic factors, reproductive history, sexual activity, HIV testing and the utilization of HIV care and treatment services, and attitudes towards HIV disclosure. Data from household and individual interviews were collected using mobile tablets and were stored on a central server [12].

All eligible participants received home-based HIV testing and counseling (HBTC) following Ethiopia’s rapid HIV diagnostic algorithm. Although participants self-reported their HIV status in the individual questionnaire, HIV status for the survey was defined by the result of the HIV rapid test. HIV rapid test results were returned to consenting participants at the household. Laboratory confirmation of seropositive samples was conducted using the Geenius HIV-1/2 supplemental assay. Self-reported awareness of HIV-positive status was confirmed by HIV-1 RNA and antiretroviral (ARV) drug testing [13].

### Variables

We assessed the demographic, behavioral, and structural factors associated with HIV testing in the year preceding the survey. We generated a three-level outcome variable comparing: 1) those who self-report testing in the 12 months preceding the survey, 2) those self-reporting testing more than 12 months since the survey, and 3) those who never tested. Those who reported ever testing for HIV and reported testing in the 12 months prior to the survey, regardless of whether they received their test results, were categorized as having tested for HIV in the last 12 months. We categorized those reporting a last HIV test date as more than 12 months preceding the survey separately. Those who did not report ever testing for HIV were categorized as having never tested.

Age, marital status, education, employment, and urban area size are covariates in this analysis. Youth were disaggregated into two five-year age groups of 15-19 years and 20-24 years. For this analysis, adolescents are persons aged 15-19 years and young adults are persons aged 20-24 years. Small and large urban areas were determined by their population size. Small urban areas were those with less than 50,000 people and large urban areas were areas with 50,000 people or more. Individuals missing data due to a response of “don’t know” or refusal to answer the questions on marital status, education, and employment were categorized with those who answered negatively to the question (never married, no education, and not employed, respectively). Those missing a response were included in the sample to maintain the overall sample size and the power within the regression models. Of this sample, 0.7% of respondents are missing or did not disclose their marital status, 0.1% of respondents did not disclose their employment status, and 0.2% of respondents are missing or did not disclose their highest level of education.

The sexual risk behaviors included in this analysis were age at first sex, the presence of a non-spousal or non-live-in sex partner in the past 12 months, condom use at last sex in the past 12 months, and engagement in transactional sex. To evaluate the accessibility of HIV testing within the Ethiopian healthcare system, we included the EPHIA variables for antenatal, tuberculosis, and sexually transmitted infection (STI) clinic attendance in our analysis.

### Data Analysis

Survey weights were applied to the responses to the household interview, the individual participant interview, and the HIV biomarker survey data. Sampling weights allow for two-cluster survey data to be generalizable beyond the sample by accounting for sample selection probability and for the nonresponse of the households and individuals included in the original sample. We analyzed responses from survey participants ages 15-24 years who had complete questionnaire and HIV testing data, and thus used the biomarker weights. Additionally, data analysis was only conducted among HIV-negative and unaware HIV-positive individuals, with awareness of HIV status confirmed through self-reporting or confirmatory ARV drug testing. Individuals that were found to have ARVs in their blood were considered aware of their HIV status and excluded from the analysis.

We generated bivariate weighted percentages and prevalence ratios (PRs) of testing for HIV in the last 12 months by select demographic, behavioral, and healthcare covariates. We conducted significance testing (p<0.05) to determine if prevalence ratios for testing for HIV in the last 12 months significantly differed from 1, and generated p-values (Table 2). We generated two multinomial logistic regression models [14], separating male and female respondents to eliminate the bias of confounding by gender. In the multinomial model, we compared the likelihood for testing in the past 12 months and testing more than 12 months before the survey to the likelihood of never testing. The models were adjusted for age, marital status, and employment in the past 12 months. To assess sexual behavioral risk for HIV infection, the variable for ‘non-spousal sex partner in the past 12 months’ was included in the final model. Similarly, medical male circumcision and antenatal care attendance served as indicators for healthcare access within their respective male and female multivariate models. We conducted significance testing to determine if the adjusted relative risk ratios significantly differed from 1, using a p-value of less than 0.05. Stata statistical software version SE 15.0 was used to perform all statistical analysis. All weighted estimates and 95% confidence intervals (CI) were calculated using jackknife replicate weights that account for sampling, nonresponse, and under coverage in the variance estimation.

### Ethics for Human Subjects Research

This study was funded by the President’s Emergency for Plan for AIDS Relief under the terms of cooperative agreement #1U2GGH001226. The study was approved by the Institutional Review Boards at the Ethiopia Public Health Institute, Columbia University Medical Center, Westat, and the Institutional Review Boards at The Centers for Disease Control and Prevention in Atlanta and Ethiopia. The survey was conducted by the Ethiopia Public Health Institute (EPHI), the Central Statistics Agency of Ethiopia, ICAP at Columbia University, Westat, and with technical assistance provided by the CDC. Participant consent was obtained through written informed consent or assent. Adult participants 18-64 years and emancipated minors 13-17 years provided informed consent prior to completing the individual interview and prior to survey blood collection. Adolescents 12-17 years were required to obtain parental permission and provide written assent to participate in the interview and blood collection.

## RESULTS

A total of 7,547 youth aged 15-24 years were interviewed from selected households and completed HIV testing (Figure 1). Of these, 0.7% (n=62) tested HIV positive; 37.2% (n=23) of those positive were confirmed to be unaware of their HIV-positive status. Our analysis examines those 7,382 youth who tested negative for HIV or were found to be unaware of their HIV-positive status and had complete HIV testing information (Figure 2). Overall, 49.6% (95% CI: 49.4-49.7, n=3,805) of all respondents were 15-19 years old and 50.5% (95% CI: 50.3-50.6, n= 3,577) were 20-24 years old, and 49.8% (95% CI: 49.6-50.0) of respondents were male (Table 1).

### Sample Characteristics

About a similar proportion of the youth participants resided in small and large urban areas (Table 1). Twenty-two percent (22.3%, 95% CI: 20.5-24.1) of all youth have ever married or lived with a partner as if married (Table 1). However, the proportion of female youth who have married was three times that of male youth. Among urban youth 15-24 years, 64% (95% CI: 61.8-66.2) report achieving secondary or post-secondary education as their highest level of education. More than two-thirds (69.4%, 95% CI: 67.5-71.3) of all youth were not employed or did not receive compensation for work in the 12 months preceding the survey (Table 1).

Nearly forty percent (39.6%, 95% CI: 37.8-41.4) of all youth 15-24 years reported ever having sex. Of these, 18.7% (95% CI: 16.5-21.0) of females and 13.4% (95% CI: 11.1-16.1) of males report sexual debut before the age of 15. Thirty-seven percent (36.8%, 95% CI: 33.5-40.3; n=682) of youth reported having at least one non-spousal or non-live-in sexual partner in the last year; but the rate was higher in males (58.1%, 95% CI: 52.4-63.5) versus females (23.0%, 95% CI: 20.2-25.9). Most (62.6%, 95% CI: 60.6-64.6) youth expressed that it is easy for them to access a condom; 72.8% (95% CI:70.0-75.5) of male youth versus 52.5% (95% CI: 50.3-54.6) of female youth. For youth who reported having sex in the 12 months preceding the survey, 83.4% (95% CI: 80.9-85.7; n=1,751) reported that they did not use a condom at last sex. Proportionally more female than male youth reported not using a condom at last sex (92.2% (95% CI: 90.6-93.6) vs. 69.3% (95% CI: 63.4-74.7), respectively).

Ninety-five percent (94.9, 95% CI: 92.9-96.3) of male youth report being circumcised; of these, 30.1% (95% CI: 26.5-33.9) had a medical circumcision (circumcision performed by a physician or clinical officer) (Table 1). Among female youth who gave birth in the three years preceding the survey, 94.5% (95% CI: 89.8-97.1) report to have received antenatal care during their most recent pregnancy (Table 1).

### Prevalence of HIV Testing in the past 12 months

Among HIV-negative and HIV-positive Ethiopian youth unaware of their HIV status, 51.4% n=3,971) report ever being tested for HIV (Figure 1). Twenty-two percent (21.8%) of these youth reported to having an HIV test in the 12 months preceding the survey and 27.5% tested for HIV more than a year before the survey (Figure 1). Proportionally more youth aged 20-24 than youth 15-19 years reported having tested in the last 12 months (31.4% (95% CI: 29.2-33.7) vs. 12.3% (95% CI: 10.9-13.8), respectively) (Table 2). Twenty-seven percent (26.8%, 95% CI: 25.2-28.6) of female youth reported testing for HIV in the last 12 months, while only 16.8% (95% CI: 14.8-19.0) of men have tested within this same timeframe.

Forty-seven (47.6%, 95% CI: 42.5-52.8) percent of female youth who attended an ANC clinic in the last three years also received an HIV test in that year (Table 2). Only 1.2% (95% CI: 0.8-1.7) of all youth reported having received a prior STD diagnosis (Table 1); of these 54.3% (95% CI: 35.4-72.1) received an HIV test (Table 2). In addition, 40.5% (95% CI: 32.8-48.8) of those who have ever been to a TB clinic also received HIV testing in the last year (Table 2).

### Predictors of HIV Testing in Youth

For all youth, age, gender, marital status, and recent employment were significant predictors of HIV testing in the 12 months preceding the survey. Youth 20-24 years of age were more than twice as likely to have tested for HIV in the last 12 months than youth 15-19 years (PR=2.6, 95% CI: 2.3-2.9). Regardless of age, women were almost twice as likely to have tested in the last year in comparison to men (PR= 1.6, 95% CI: 1.4-1.8). Those ever married or have lived with a partner were three times as likely to have tested in the last 12 months (PR=2.8, 95% CI: 2.5-3.1). Employed youth were nearly twice as likely to have HIV tested in the past year than unemployed youth (PR=1.6, 95% CI: 1.4-1.8). For all youth, the likelihood of HIV testing in the last 12 months did not significantly increase by level of education (Table 2).

Youth who have been sexually active (PR=3.2, 95% CI:2.8-3.6), and those who received an STD diagnosis in the last year (PR= 1.5, 95% CI: 1.0-2.1) were significantly more likely to have tested for HIV in the last year than to have never tested (Table 2). Those reporting at least one non-spousal, non-live in sexual partner in the past 12 months were significantly less likely to have tested for HIV in the past year, in comparison to those with all marital or live-in partners (PR=0.74, 95% CI: 0.6-0.9). Youth who report using a condom at last sex were significantly less likely to have tested for HIV in the last year (PR=0.80, 95% CI: 0.6-1.0).

The youth in this analysis were significantly more likely to test for HIV in the past 12 months if they had ever visited a TB clinic or received care for STI symptoms (Table 2). Males who had medical male circumcision were significantly less likely to have tested for HIV in the past year (PR=0.70, 95% CI:0.5-0.9).

Table 3 presents the multinomial logistic regression models for HIV testing by gender. Among males, controlling for variables of sexual behavioral risk and healthcare access, being older (20-24 years) was the only significant predictor for recent HIV testing likelihood. Young adult males (20-24 years) in comparison to adolescent males (15-19 years), had a three-fold likelihood of testing for HIV in the past year (RR=2.8, 95% CI:1.4-5.7) than never testing. Males who reported a non-spousal, non-live in sexual partner in the last year were significantly less likely to have tested for HIV in the last 12 months (RR=0.3, 95% CI:0.1-0.8). When controlling for other variables, marital status did not have a significant effect on the likelihood for receiving a recent HIV test among men in this age group (Table 3). For non-recent testing (≥12 months before the survey), young adult males (20-24 years) were significantly more likely to have ever tested in comparison to adolescent males 15-19 years (RR=3.0, 95% CI: 2.0-5.5). There was not a significant difference between men reporting that all their partners in the last 12 months were marital partners and men reporting no marital partners and their likelihood to have ever tested for HIV (RR=0.8, 95% CI:0.3-2.0).

Among female youth, recent HIV testing (<12 months) increased with older age (20-24 years old), having ever married or lived with a partner, and having at least secondary education (Table 3). Also, females who attended an antenatal clinic in the last three years were significantly more likely to have tested for HIV in the last year (Table 3). When controlling for factors such as age or education level, female youth who reported a non-spousal, non-live-in sex partner in the past 12 months, were not significantly more likely to have HIV testing, either within the last year or more than one year before the survey (Table 3).

## DISCUSSION

Among youth ages 15-24 in urban Ethiopia, the individual factors that predict an increased likelihood for testing for HIV in the last year is female gender, older age (20-24 years), and ever being married. Looking at sexual risk behaviors, 40% of all youth report ever having sex and 40% of whom report engaging in sex with a non-spousal, non-live-in partner in the last year. Youth reporting to have had a non-marital sex partner in the last year were less likely to have recently tested for HIV by a factor of 0.64, in comparison to those who report only marital sex partners. Screening for TB infection or a sexually transmitted disease each increases the likelihood for testing in the last year by two-fold. Male youth who received a medical circumcision were significantly less likely to be HIV tested in the last 12 months compared to those who did not have a medical circumcision. When these factors were modelled together, older age (20-24 years), continues to be a significant factor in predicting current engagement in HIV testing for both male and female youth. Sexual risk behaviors are an important factor to consider in predicting recent testing behaviors among males; and marital status and healthcare access remain persistent predictors for female youths’ access to current HIV testing.

Our findings are consistent with the Ethiopian Federal Ministry of Health HIV Counseling and Testing (HCT) Guidelines [10]. Women and those who have ever married are more likely to have tested for HIV in the past year, as national guidelines emphasize the promotion of HIV counseling and testing to couples and women. Couples are encouraged to test at ‘pre-engagement, pre-marital, or pre-conception’ stages, with partner notification services offered. While partner testing should be encouraged, marital status is not a significant predictor of HIV testing in the last year in male youth. Recent data from the Demographic Health Survey (DHS) shows that Ethiopian male youth are likely to sexually debut 2.5 years before entering marriage [7], with urban males marrying even later. As adolescent boys reach young adulthood, undiagnosed HIV infection may contribute to HIV transmission.

National guidelines also promote testing to women during pregnancy and at the time of labor [10]. Additionally, guidelines encourage the integration of HIV counseling and testing within other health services, including TB and STI screening. The age of sexual consent in Ethiopia is eighteen years and, according to the national guidelines, adolescents may consent to HIV counseling and testing services at the age of 15 without approval from a parent or guardian [10] [15]. Youth aged 13-15 years old who are sexually active (through the commercial sexual exploitation of minors or child marriage) are considered ‘mature minors’ and can provide informed consent without parental permission [10]. Within our sample, eight percent of female youth and five percent of male youth reported to have engaged in sex before the age of 15 (Table 1), well before the legal age of consent. It is possible that youth who have engaged in sexual activity before the legal age are less likely to seek consistent testing due to stigma and societal expectations.

The 2007 National Guidelines for HIV Counseling and Testing in Ethiopia encourages the availability of ‘youth friendly services’, but these services are not clearly defined or linked to an existing health program [10]. Improving the comprehensive knowledge of HIV among youth, specifically among male youth, may be an effective strategy to increase HIV testing in this age group. An analysis of DHS data from 29 sub-Saharan African countries found an association between comprehensive HIV education and HIV testing in men [17]. Countries like Rwanda have found that HIV education disseminated through newspaper, radio, and television media campaigns were effective at reaching males and youth [17]. In Zambia and South Africa, in communities that achieved 90% awareness of HIV status among female youth, the door-to-door distribution of home-based self-test kits in select communities, proved to be effective at increasing the update of HIV testing and awareness of HIV status among males 16-29 years [18]. In Malawi, where HIV prevalence in youth is relatively low in comparison to adults, an index-testing strategy to test the children of HIV-positive adults through home-based testing yielded a 94% uptake of HIV testing among children <15 years and youth 15-24 years [19].

The results of our analysis uniquely differ from much of previous research on adolescent and young adult HIV testing in Africa. In an analysis of DHS data from four sub-Saharan African countries, males and those ages 15-19 years were less likely to have tested for HIV in comparison to their older, female counterparts [16]. Our inclusion of adolescent boys and young men, 15-24, highlight an additional demographic for which HIV testing services should be targeted. Additionally, our analysis examines the engagement of HIV testing within the last 12 months. Examining HIV testing with this temporal restriction allows for a more accurate evaluation of the reach of current HIV testing and counseling strategies.

### Limitations

The implications of our analysis are limited by the nature in which the data were collected. As a household survey, HIV testing history was captured through self-report rather than from an electronic medical record. Our data is subject to recall bias as respondents may have forgotten if they have ever tested or misreported the date of their last HIV test. The data is also subject to social desirability bias. The age of sexual consent in Ethiopia is eighteen [14]. Youth younger than 18 years of age may not disclose their prior sexual activity or engagement in HIV testing services to avoid any cultural stigma associated with engaging in sex before the age of consent. Estimates for testing for HIV and sexual risk behavior are likely underreported as a result. Lastly, the inclusion of unemancipated minors (15-17 years) in our sample required that they be related to an eligible and consenting adult. Our findings may better characterize the HIV testing behaviors of emancipated minors, 15-17 years old. Emancipation status was not included as a possible predictor of testing due to our sample including youth who are and are not of legal age.

## Conclusion

In urban Ethiopia, age, gender, and marital status significantly predict the engagement of youth in recent HIV testing. These predictors show that adolescents, aged 15-19 years, and males are not being adequately reached within the national HIV counseling and testing strategy in urban Ethiopia. Adolescent girls have a high probability of obtaining testing services if pregnant, through the promotion of HIV testing services in antenatal care. Our findings show that adolescent males are likely to be left out of the existing Ethiopia’s targeted HIV testing strategy.

Among 15-24 years old males who report recent sex, most report sex outside of a committed partnership and few report consistent use of HIV prevention methods. Efforts to increase comprehensive knowledge of HIV and expand access to HIV testing, through at-home, self-testing kits, have been proven to be effective at reaching adolescent males in national HIV testing efforts. Expanding current HIV testing efforts to include male youth friendly services is important for the future of HIV epidemic control in urban Ethiopia.

## Data Availability

The 2017-2018 EPHIA public release datasets and files are available at the ICAP PHIA website (https://phia-data.icap.columbia.edu/datasets?country_id=12). Survey data use manuals and documentation are available for download. For access to datasets, please register for an account and submit a data request form.

## ACKNOWLEDGEMENTS

We extend our thanks to the survey investigators and contributors from the Federal Ministry of Health, EPHI, ICAP, Westat, and CDC for their leadership and coordination of the survey. We also thank and acknowledge the EPHIA study group, specifically the data collection staffs, the field teams, laboratory, and administrative staff for their hard work and dedication to complete the survey. We also thank all EPHIA study participants for their gracious participation.

